# *TP53* Variant Clusters Stratify the Li-Fraumeni Spectrum and Reveal an Osteosarcoma-Prone Subgroup

**DOI:** 10.1101/2024.01.06.23300162

**Authors:** Nicholas W Fischer, Brianne Laverty, Noa Alon, Emilie Montellier, Kara N Maxwell, Christian P Kratz, Pierre Hainaut, Ran Kafri, David Malkin

## Abstract

Li-Fraumeni syndrome (LFS) has recently been redefined as a ‘spectrum’ cancer predisposition disorder to reflect its broad phenotypic heterogeneity. The wide functional gradient associated with different *TP53* variants is thought to contribute to LFS heterogeneity, although it is still poorly understood and there is an unmet clinical need for risk stratification strategies. Leveraging p53 mutagenesis dataset, we performed an unsupervised cluster analysis that revealed five *TP53* variant clusters with unique structural and functional consequences. Classifying variant carriers according to these clusters stratified cancer onset and survival using discovery and validation cohorts, and exposed important clinical characteristics to consider for patient management. In particular, we identified a subgroup of monomeric *TP53* variant carriers prone to osteosarcoma, along with a cluster associated with less “LFS-like” phenotypes enriched in carriers with no history of cancer. Our classification of *TP53* variants demonstrates the existence of a wide *TP53-*heritable cancer susceptibility spectrum and provides a new framework to delineate carriers toward personalized patient care.

## INTRODUCTION

Inherited cancer susceptibility in Li-Fraumeni Syndrome (LFS; MIM #151623) was first described in 1969 based on the observation of several families with a high incidence of rare and diverse cancer types [1]. Since this initial observation, germline *TP53* mutations were identified as the underlying cause of LFS [2, 3] and the clinical classification has been adapted by multiple revised sets of criteria [4–7] to incorporate individuals who do not fit the strict ‘classic LFS’ diagnosis [8]. Recently, LFS was reclassified as a ‘spectrum’ of disease rather than a syndrome to reflect the heterogeneity that has become increasingly evident through genotype-phenotype association studies [9]. The LFS spectrum encompasses a highly variable age-of-onset with nearly 30% affected during childhood, largely unpredictable sites of tumor manifestation, and inconsistent cancer penetrance. To date, the factors causing this heterogeneity in LFS remain unclear, although unequal mutant p53 functional consequences have been linked to different cancer phenotypes [10–12].

Increased use of DNA sequencing in the clinic has led to the discovery of hundreds of novel germline variants that span the entire *TP53* gene. Missense mutations leading to single amino acid changes represent the vast majority of *TP53* variants that are associated with LFS. There are thousands of possible single amino acid substitutions in p53, and currently, at least 1,703 different germline variants have been linked to individuals with cancer, as catalogued by the National Cancer Institute’s (NCI) *TP53* Database (https://tp53.isb-cgc.org). However, research efforts have focused on variants at a few “hotspot” residues that account for only 30% of germline *TP53* missense variants found in patients, leaving the majority largely uncharacterized and many still considered variants of uncertain significance (VUS). Therefore, it is of great interest to classify all variants in order to provide more accurate cancer risk assessments with tailored surveillance strategies and treatment plans.

Mutagenesis studies are an important clinical tool used to assess *TP53* variant pathogenicity, however, it is often unclear without substantial clinical evidence that is difficult to obtain from predominantly rare and infrequent variants. The results from large-scale mutagenesis experiments have demonstrated a broad range of functional consequences associated with different variants, ranging from near wild-type (WT) p53 capabilities to severe loss-of-function (LOF) [13–15]. As a multifaceted protein, p53 is composed of several domains containing complex structured and unstructured regions that enable a myriad of cellular tasks. Primarily operating as a transcription factor, p53 binds to DNA recognition sequences as a tetramer and regulates the expression of several thousand target genes to orchestrate cellular responses centering around DNA-damage repair (DDR) and cell fate decisions [16]. Hence, mutations arising at different locations of the protein can impart different cellular consequences by altering its transcriptional activity (TA) as well as its non-TA. Some variant p53 species can also exert a dominant-negative (DN) effect over WT p53 [17, 18]. This is an important feature because LFS and heritable *TP53*-related cancer predisposition follows an autosomal dominant inheritance pattern and most patients are heterozygous carriers who retain one copy of WT *TP53*. Moreover, mutant p53 LOF can vary under different cellular conditions. For example, the R337H variant—the most frequently reported germline variant due to a genetic founder event in Brazil—has a conditional pH-dependent LOF and is associated with the tissue-specific development of adrenocortical carcinoma (ACC) [19, 20]. Thus, the functional assessments derived from comprehensive variant library screens conducted in varying cellular contexts provide clinically relevant measurements of specific p53 activities.

Here, we incorporated p53 variant library screens using an unsupervised cluster analysis and uncovered five functionally distinct variant clusters. When applied to germline variant carriers from the NCI *TP53* Database and a validation dataset, we identified unique cancer patterns and significant differences between ages at onset and cancer survival. Our variant clustering model provides a new classification tool for variant interpretation and risk stratification of *TP53* variant carriers.

## METHODS

### Functional datasets and unsupervised cluster analysis

We selected saturation mutagenesis cellular functional screens as features for dimensionality reduction. Giacomelli et al. measured p53 cellular loss-of-function (LOF; nutlin-3 treatment in p53^NULL^ human A549 cells), dominant-negtive activity (DN; nutlin-3 treatment in isogenic p53^WT^ A549 cells), and DNA-damage repair (DDR; etoposide treatment of p53^NULL^ A549 cells) using competitive growth assays [14]. Kato et al. measured transcriptional activity (TA) in yeast using a transcriptional reporter assay [13]. We used the median TA values (8 p53 promoter-specific elements) normalized to WT p53. Matrix correlation plots comparing scaled data were generated using *pairplot* in Python’s Data Visualization library *Seaborn* using the R package *reticulate*. Dimensionality reduction was performed using PCA (principal component analysis) and UMAP (Uniform Manifold Approximation and Projection) with R packages *ggfortify* and *umap*. The R package *cluster* was used to determine the number of clusters to be generated (Gap statistic, elbow method, and silhouette score) and k-means clustering. Data was visualized using the R package *ggplot2*.

### Structure/function analysis

Rendering of the structure of two p53 core DNA-binding domains bound to DNA (PDB #3EXJ) was done using PyMOL software. Structural/functional domain annotations for each variant were obtained from the NCI *TP53* Database. Prediction of intrinsically disordered regions was conducted using IUPred3 [21].

### Clinical datasets, curation, and selection criteria

The National Cancer Institute’s (NCI) *TP53* Database (version R20; https://tp53.isb-cgc.org) is a repository for germline variant carriers that was used as the discovery cohort (n=3,446) to evaluate clinical associations. Carriers of multiple variants were removed from the dataset because the relative contribution of the variants is unknown. In addition, carriers of the R337H variant (n=291) were removed from the dataset due to a known sampling bias as a founder variant in Brazil. Patients or tumors with no ID were removed to avoid inclusion of duplicates. The validation dataset (n=458) is a multi-institutional collection of individuals and families carrying germline *TP53* variants from registries at three cancer centers: in Canada (The Hospital for Sick Children), the United States (Perelman School of Medicine at the University of Pennsylvania), and Germany (Hannover Medical School). The cohort from Germany included only carriers of pathogenic or likely pathogenic variants based on the Fortuno classification [22]. All carriers of protein-truncating variants (frameshift, non-sense, and deletions) were included as a separate group (FS/NS/DEL) in each analysis.

### Comparisons with non-cancer associated datasets and the Variant Curation Expert Panel (VCEP) annotations

We used the Genome Aggregation Database (gnomAD; https://gnomad.broadinstitute.org/) v3.1.2 non-cancer dataset mapped against the canonical transcript (Ensembl transcript ID ENST00000269305.9) to investigate the prevalence of variants in the general population with no history of cancer. The Fabulous Ladies Over Seventy (FLOSSIES) Database (https://whi.color.com/) contains germline DNA sequencing from 27 genes (including *TP53*) collected from 10,000 women over 70 years of age who have never had cancer. We identified 27 *TP53* variants in this dataset and mapped them against the *TP53* clusters. *TP53* variant clusters were also compared to VCEP annotations, which were obtained from the ClinVar database (https://www.ncbi.nlm.nih.gov/clinvar/).

### Survival analysis

The lifetime survival analysis was performed using clinical data obtained from the NCI *TP53* Database and the validation dataset. Unaffected carriers were included as censored events. The 10-year cancer survival analysis was determined using age at diagnosis to age at death. Patients were censored at time of last follow-up if not deceased. The R package *survival* was used to compute Kaplan-Meier (KM) estimates and hazard ratios, and the *survminer* package was used to generate KM plots.

### Statistical analysis

All statistical tests were performed using R or GraphPad Prism (v9). Linear relationships in matrix correlation plots were assessed using Pearson’s correlation coefficient. Kruskal-Wallis tests were used to determine differences between functional scores, Grantham’s distance, and ages at onset across all groups. Pairwise comparisons were then performed using Mann-Whitney U tests. Chi-squared tests were used to determine significance across groups for cancer type comparisons in variant carriers and differences of proline substitutions in clusters. Subsequently, pairwise comparisons were done using Fisher’s exact test. All multiple hypothesis corrections for pairwise tests were done using the Benjamini-Hochberg adjustment to obtain the false discovery rate (FDR). KM survival estimates were compared using log-rank tests. Hazard ratios (HR) with 95% confidence intervals (CIs) were calculated using the Cox proportional hazards regression analysis.

## RESULTS

### Unsupervised learning identifies five clusters of TP53 variants based on p53 cellular functional features

Various functional features of p53 have been evaluated using *TP53* saturation mutagenesis screens conducted with different cellular systems and drug treatments. These features include TA, DN activity, LOF, DDR, and proliferation [13–15]. Each study has independently reported a wide-ranging functional gradient using separate variant libraries. In a preliminary analysis, we compared the results of these large-scale assays and observed weak to moderately strong inter-assay correlations (**Fig. 1a** and **Supplementary Fig. 1a**). We found that the correlations were influenced by the variant locations by examining the assay results in relation to the resident functional domains (**Fig. 1a** and **Supplementary Fig. 2a**). Strong correlations were observed between variants within the DNA-binding domain (DBD), whereas these relationships were weak or absent for C-terminal domain (CTD) variants. N-terminal domain (NTD) and oligomerization domain (OD) variants had weak to moderate linear relationships when comparing between LOF, DDR, and TA, but did not exhibit DN activity, which was almost exclusively found in DBD variants.

**Figure 1.**
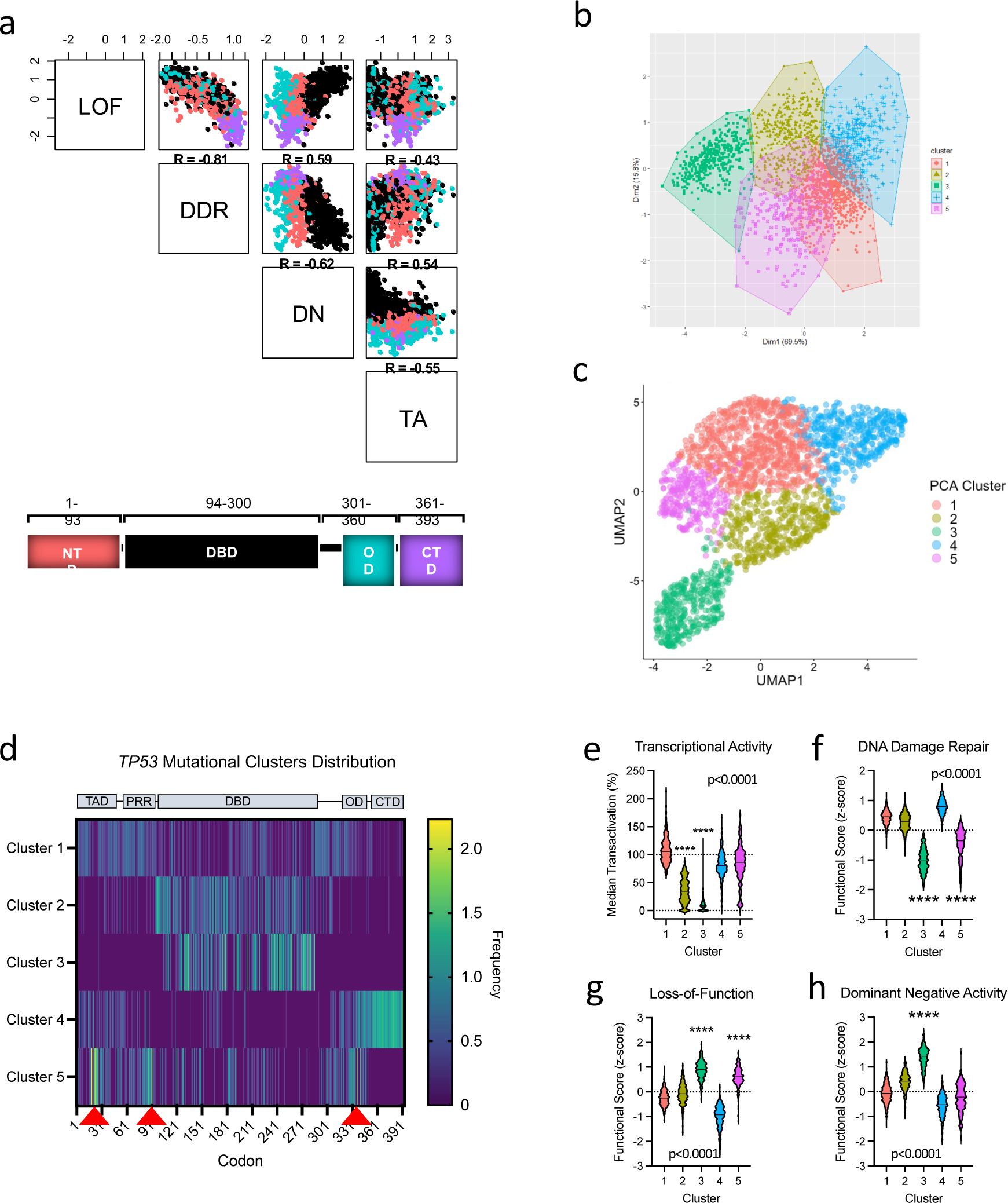
Unsupervised cluster analysis of *TP53* missense variants based on cellular p53 functional features. **(a)** Matrix correlation plots comparing functional datasets from p53 mutagenesis datasets covering the entire protein (LOF = loss-of-function; DDR = DNA damage repair; DN = dominant negative activity; TA = transcriptional activity). Variants are colour-coded based on the functional domain they reside in (NTD = N-terminal domain; PRR = proline-rich region; DBD = DNA-binding domain; OD = oligomerization domain; CTD = C-terminal domain). **(b)** Principal component analysis (PCA) and unsupervised k-means clustering performed with p53 mutagenesis cellular functional assay measurements. **(c)** Uniform manifold approximation and projection (UMAP) performed using p53 mutagenesis cellular functional assay measurements and colour-coded based on PCA k-means clustering (N=200, D=0.4). **(d)** Heatmap displaying the codon frequencies and distributions of *TP53* variant clusters. Red arrowheads indicate mutational hotspots in cluster 5. **(e-h)** Violin plots displaying the functional consequences of variants within each cluster (dotted line represents wild-type p53). P-values on plots were calculated using Kruskal-Wallis tests. Mann-Whitney U tests were used for pairwise comparisons (****p<0.0001).

We employed principal component analysis (PCA) for visualization and dimensionality reduction of the functional features measured by mutagenesis screens covering the p53 protein. At least two spatially separated clusters were visible within a clearly disjointed distribution of data points (**Supplementary Fig. 1b**). Variants located in the same domain demonstrated a tendency to cluster together, with minimal spatial separation between domains, except for a distinct subgroup of DBD variants (**Supplementary Fig. 2b**). Notably, each of the cancer-associated hotspot variants as well as monomeric p53 variants were grouped in close proximity, respectively (**Supplementary Fig. 2b**). Next, unsupervised k-means clustering was performed with five predicted clusters (**Fig. 1b**) as determined by the gap statistical method (**Supplementary Fig. 1c**). To validate this with a non-linear approach, we applied uniform manifold approximation and projection (UMAP) which demonstrated congruent results to the PCA with a similar spatial distribution of data points with one clearly isolated cluster of DBD variants (**Fig. 1c**).

### TP53 variant clusters have distinct signatures affecting different structural and functional characteristics

By mapping the clusters onto the p53 coding sequence, we identified unique variant signatures (**Fig. 1d**). First, variants in cluster 1 were found throughout the protein and broadly coincide with intrinsically disordered regions predicted by the IUPred3 computational method [21] (**Fig. 1d** and **Supplementary Fig. 3a**). In contrast, clusters 2 and 3 were centered within the highly structured DBD. Cluster 3 variants were particularly enriched within helix 2 (H2) of the loop-sheet-helix motif (L1/S/H2) that binds to the major groove of DNA, whereas variants in cluster 2 were primarily located in regions distal to the DNA contact sites (**Supplementary Fig. 3b, c**). Cluster 4 variants were distinctively concentrated at the CTD (**Fig. 1d**). Lastly, cluster 5 has a wide variant distribution with three sites of markedly increased variant frequencies located in structured regions known to impact protein stability (**Fig. 1d**). These three loci were found within TAD1 (transactivation domain 1), OD, and an NTD hinge region (residues 86-93) associated with stabilizing the p53 tetramer through its interaction with the DBD [23]. Specifically, the residues affected most frequently in cluster 5 are involved in binding to the main p53 negative regulator MDM2 (residues F19, W23, and L26) [24], DBD stabilization (W91) [23], p53 oligomerization (I332, R337, and F341), and transactivation (F19, L22, W23, and L26) [25]. We found a significant difference between the expected frequency of proline substitutions, which tend to be more structurally disruptive residues, when comparing between the clusters (p<0.0001, chi-squared test). Cluster 5 contains the most substitutions to/from proline, even after the removal of the proline-rich region (PRR; residues 60-90) (**Supplementary Fig. 3d**). Clusters 5 and 3 both contain a higher proportion of substitutions to proline compared to clusters 1, 2, and 4 (**Supplementary Fig. 3e**).

Substitutions in each cluster were assessed using Grantham’s distance, which predicts the evolutionary distance between amino acid substitutions, where higher scores are considered more deleterious. Although cluster 5 contains a higher frequency of proline substitutions, cluster 3 has the highest overall Grantham score compared to all other variant clusters (p<0.0001, Mann-Whitney U test, **Supplementary Fig. 3f**). Collectively, cluster 3 variants have the most severe functional consequences in cellular assays, including a high degree of DN activity (**Fig. 2e-h**). In contrast, cluster 1 variants are hypermorphic and cluster 4 primarily consists of variants with near-WT functionality. Cluster 2 variants are hypomorphic, exhibiting partial TA, WT-like DDR, and slight DN potential. Lastly, cluster 5 variants exhibit high LOF and impaired DDR, however, most variants in this cluster retain TA and lack DN activity.

**Figure 2.**
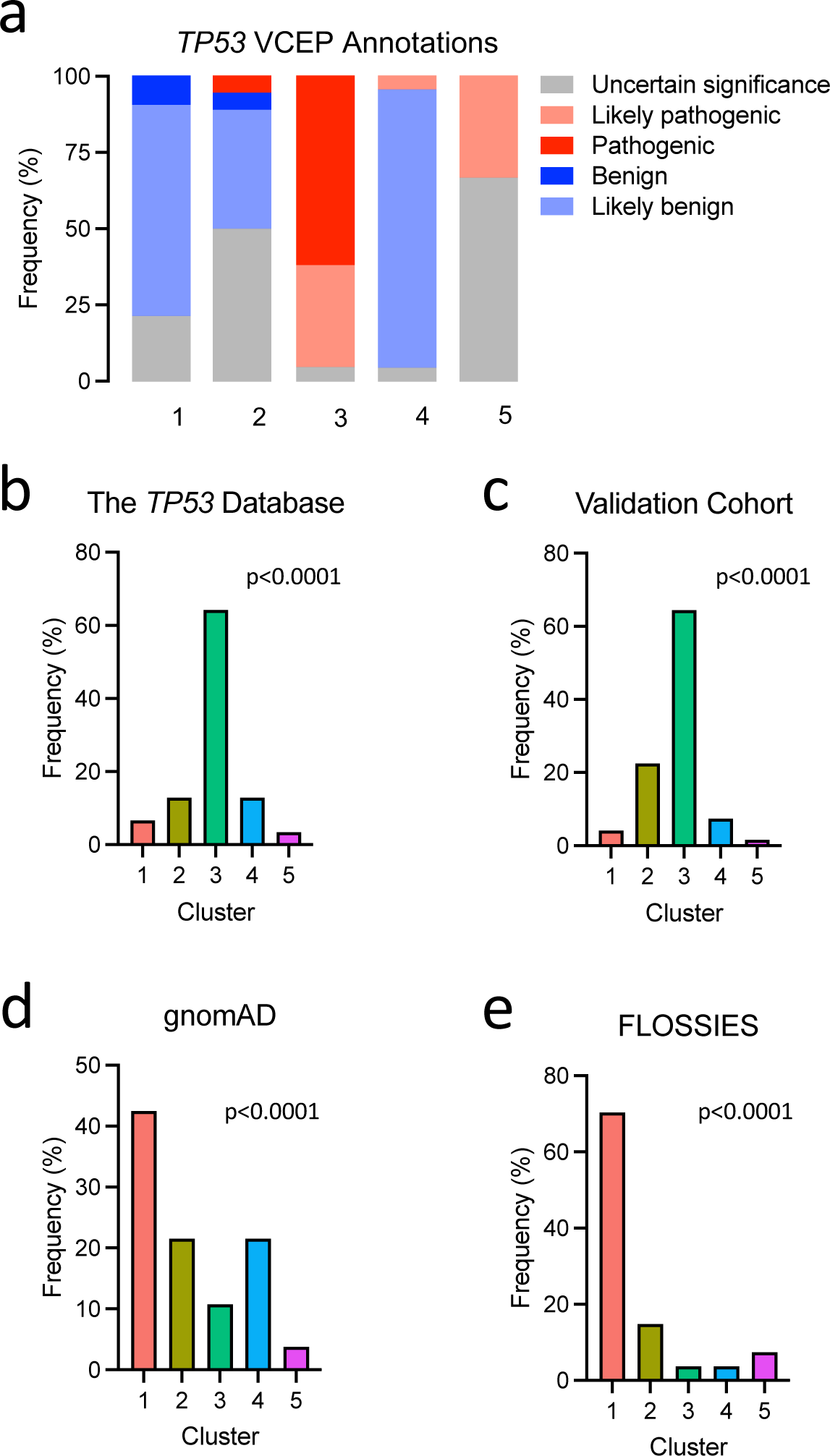
Alignment of *TP53* clusters with the Variant Curation Expert Panel (VCEP) annotations and the representation of clusters across populations with and without cancer. **(a)** Stacked bar plot showing VCEP annotated variants (n=109) applied to the variant clusters. **(b)** Bar charts displaying the frequencies of germline variants in each cluster found in the NCI *TP53* Database (n=3,113), **(c)** a multi-institutional validation cohort (n=458), **(d)** gnomAD non-cancer dataset (n=186), and (e) FLOSSIES database (n=27). Pvalues on plots were calculated using chi-square tests.

### TP53 variant clusters are clinically relevant and display differing associations with cancer

The *TP53* Variant Curation Expert Panel (VCEP) plays an important role in evaluating molecular and clinical evidence to provide variant interpretations accessible through ClinVar (https://clinicalgenome.org/affiliation/50013) [26]. VCEP uses variant functional information, including the mutagenesis assays used in our variant clustering model, in addition to clinical and population data to determine variant pathogenicity. However, *TP53* variants can be exceedingly rare and often lack sufficient clinical evidence to establish a clear association with cancer—particularly those with reduced penetrance or adult onset [27]. Thus, despite great efforts, only 109 of 2,314 potential missense variants within the gene have undergone evaluation, and still, 24 of these remain VUS. We compared the 109 VCEP annotated variants to our unsupervised clustering model to examine the clinical relevance of each variant cluster (**Fig. 2a**). A remarkable division was observed between pathogenic/likely pathogenic (P/LP) and benign/likely benign (B/LB), with cluster 3 displaying a complete separation for P/LP and cluster 1 for B/LB. Using our strategy, variant interpretations of P/LP and B/LB could be extended to over 1100 variants classified in clusters 1 and 3. Within clusters 2, 4, and 5, the distribution of VCEP annotated variants were mixed or contained more VUS. If *TP53* variant pathogenicity is a gradient resulting in a spectrum of cancer risk, then clusters 2, 4, and 5 may represent intermediate pathogenicity between B/LB cluster 1 and P/LP cluster 3. Subsequently, we explored germline sequencing datasets to determine the prevalence of each variant cluster in individuals with or without a history of cancer (**Supplementary Table 1**). Cancer-associated germline variants in the NCI *TP53* Database were predominantly classified in cluster 3 (1998/3,113 or 64%) (**Fig. 2b**). Similarly, cluster 3 variants also comprised over 64% of those identified in a multi-institutional validation dataset (n=458) collected from cancer centers across Canada, the United States, and Germany (**Fig. 2c**). Non-cancer associated germline variants from the gnomAD database, representing the general healthy population, were most often classified in cluster 1 (79/186 or 42.5%) (**Fig. 2d**). Moreover, cluster 1 variants were highly enriched in healthy older women >70 years (19/27 or 70%) with no history of cancer (FLOSSIES database) (**Fig. 2e**). Overall, the divergence of variant clusters in germline carriers affected versus unaffected by cancer and the alignment with VCEP annotations supports the potential value of our clustering strategy to delineate germline carriers into risk stratified groups.

### Classification of TP53 variant carriers based on clusters reveals unique cancer-type distributions and an osteosarcoma-prone subgroup

Individuals carrying a pathogenic germline *TP53* variant are affected by a broad range of cancers during childhood and adulthood. We classified *TP53* variant carriers in the NCI *TP53* Database based on the five clusters and investigated the cancer-type distributions in each group. Carriers of protein-truncating variants (frameshift, nonsense, and deletions; FS/NS/DEL) were included as another variant group. Each group of patients exhibited a diverse range of cancers (**Fig. 3a**), however, our analysis revealed significant differences (chi-squared test) between the occurrence of specific cancer types when comparing across all groups (**Supplementary Table 2**). Strikingly, osteosarcomas occurred at more than twice the frequency in patients carrying a cluster 5 variant (15.8%; 16/101 patients) compared to all other cluster-based groups (p<0.0001, chi-squared test) (**Fig. 3b** and **Supplementary Table 3**). Pairwise comparisons determined that the odds of developing osteosarcoma for cluster 5 variant carriers is significantly higher than in all other cluster-based patient groups, with odds ratios of (OR)=6.3 versus cluster 1 (95% confidence interval (CI)=2.1-22.8; FDR=0.0015, Fisher’s exact test), OR=4.6 versus cluster 2 (95% CI=2.0-10.6; FDR=0.0015, Fisher’s exact test), OR=2.5 versus cluster 3 (95% CI=1.3-4.4; FDR=0.0092, Fisher’s exact test), and OR=14.5 versus cluster 4 (95% CI=2.2-622; FDR=0.0029, Fisher’s exact test) (**Supplementary Table 4**). This association did not reach significance when comparing cluster 5 to FS/NS/DEL carriers in which the frequency of osteosarcoma was 9.0% (71/788 patients) (OR=1.9; 95% CI=0.98-3.5; FDR=0.065, Fisher’s exact test). All bone tumors in cluster 5 (n=17) were classified as osteosarcomas with the exception of one unclassified bone sarcoma (not otherwise specified). Notably, upon analysis of the *TP53* variants in cluster 5 patients, we found that the majority were located in the OD and known to abolish oligomerization, resulting in monomeric p53 species (L344P, R337C, R337L, R337P, and R342P, in 81 of 88 individuals from 20 unrelated families) [12, 28]. For carriers of a monomeric p53 variant, osteosarcomas represented 16.8% (16/95 patients) of all cancers developed in this subgroup (**Supplementary Table 3**).

**Figure 3.**
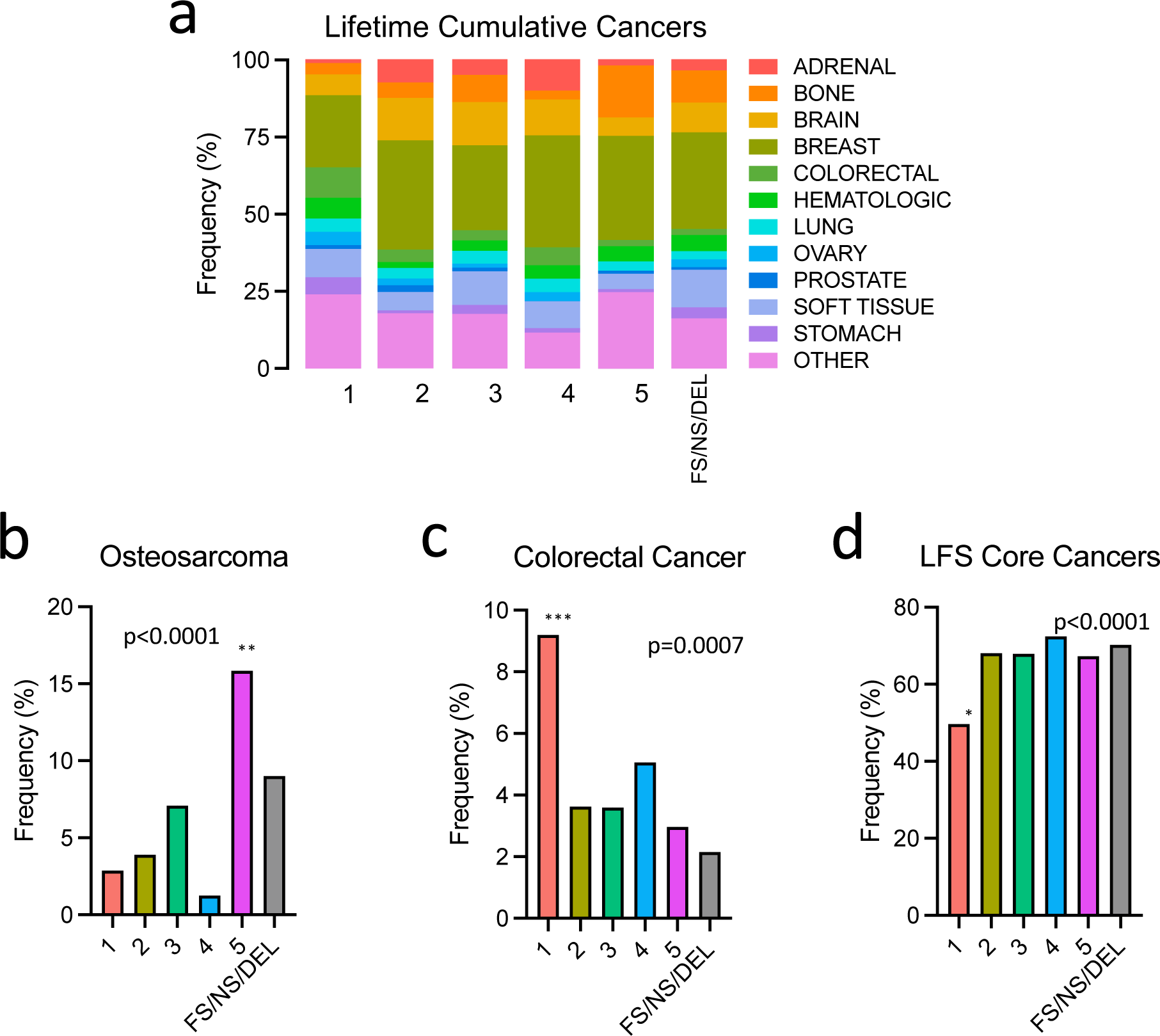
Cancer spectrums in germline *TP53* variant carriers classified based on the variant clusters. **(a)** Stacked bar plot displaying the cancer type distribution patterns manifested in germline *TP53* mutation carriers. **(b)** Bar charts showing the frequency of osteosarcoma, **(c)** colorectal cancer, **(d)** and LFS-core cancer types according to the cluster-based patient classification. Pvalues on plots were calculated using chi-square tests. Fisher’s exact tests were used for pairwise comparisons (*p<0.05, **p<0.01, ***p<0.001).

Next, we observed a greater proportion of colorectal cancers in cluster 1 variant carriers. Colorectal cancer is considered relatively uncommon in LFS, representing 3.4% (136/3987) of total cancers in NCI *TP53* Database. However, we found a significant difference between the patient groups (p=0.0007, chi-squared test), in which cluster 1 carriers exhibited the highest frequency of colorectal cancer affecting 9.2% (16/174) of patients (**Fig. 3c** and **Supplementary Table 5**). In addition, we identified a significant difference between the expected frequencies of the core cancers that are commonly associated with LFS: osteosarcoma, soft tissue sarcomas (STS), breast tumors, brain tumors, adrenocortical carcinoma (ACC), and leukemia (p<0.0001, chi-squared test) (**Supplementary Table 6**). Interestingly, carriers of cluster 1 variants presented with considerably fewer LFS core cancers (49.7%) versus all other patient groups which each totalled approximately 70% LFS core cancers (**Fig. 3d**). The odds of manifesting an LFS core tumor for cluster 1 carriers was at least 2.1-fold lower than all other groups (FDR<0.015, Fisher’s exact test) (**Supplementary Table 4**). Together, these observations of unique tumor patterns suggest that *TP53* variant structural and functional features can impact cancer pathogenesis in germline carriers.

### TP53 variant clusters significantly stratify cancer onset and survival of carriers

Carriers of *TP53* variants endure a lifelong high susceptibility to cancer with wide-ranging ages at onset, and some experience multiple cancers. After separating *TP53* variant carriers into cluster-based groups, we found significant differences between the ages at diagnosis of cancer (p<0.0001, Kruskal-Wallis test) (**Fig. 4a,b**). Cancer diagnoses were significantly earlier in carriers of clusters 3, 5, and FS/NS/DEL variants when compared to carriers of variants in clusters 1, 2, and 4, with median age differences of at least 7 years (**Fig. 4a** and **Supplementary Table 7**). Importantly, our validation cohort demonstrated the same pattern of divergent ages at diagnosis (p<0.0001, Kruskal-Wallis test), where cluster 1 carriers exhibited the oldest onset, while clusters 3, 5, and FS/NS/DEL carriers consistently displayed the earliest onset (**Fig. 4b** and **Supplementary Table 7**).

**Figure 4.**
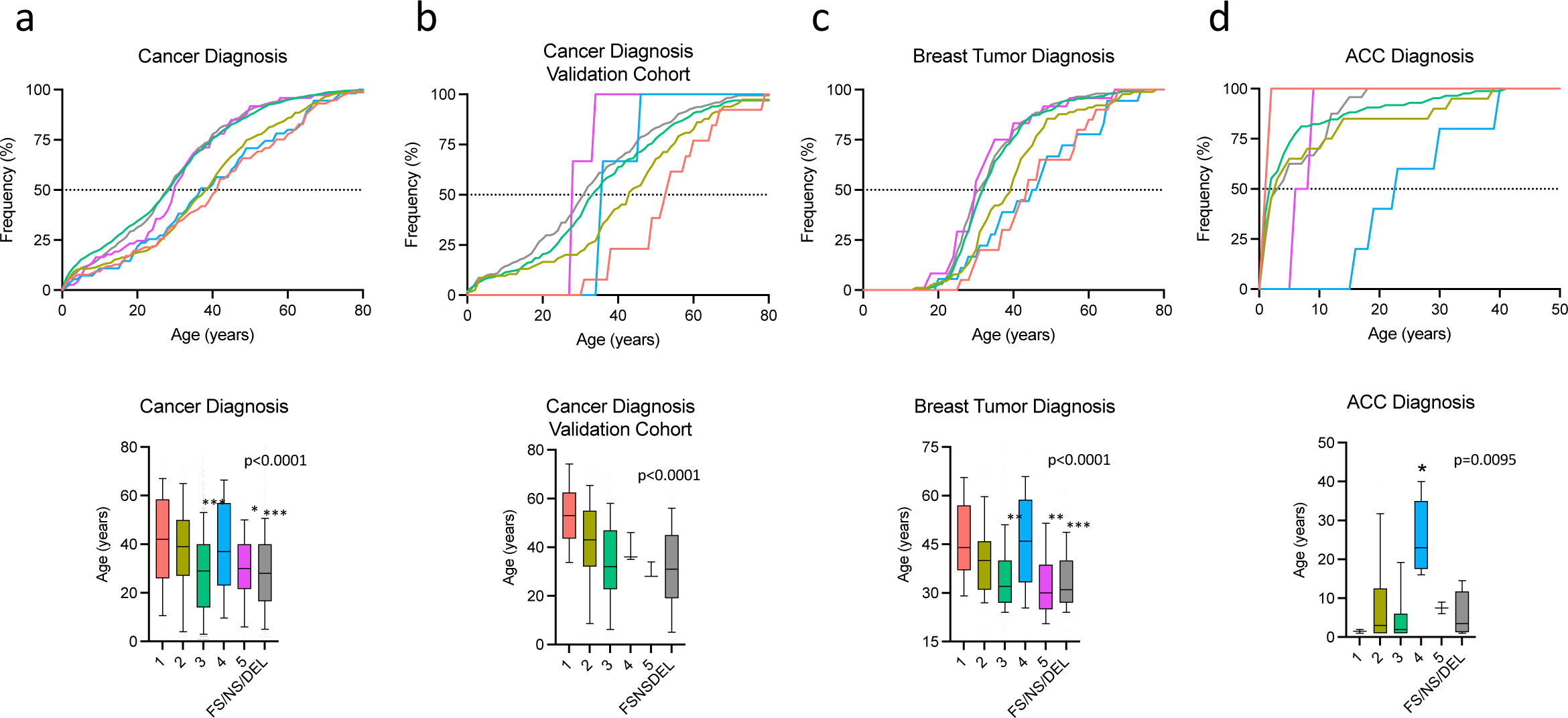
Age at cancer diagnosis in germline *TP53* variant carriers. **(a)** Line plot displaying the cumulative cancer diagnosis ages over time and a box and whiskers plot (10-90^th^ percentiles) representing the ages at cancer diagnosis in the NCI *TP53* Database, and **(b)** in the validation cohort. *compared to clusters 1, 2, and 4. **(c)** Line plot displaying the frequency of breast tumor diagnosis over time and a box and whiskers plot (10-90^th^ percentiles) representing the ages at cancer diagnosis. *compared to clusters 1, 2, and 4. **(d)** Line plot displaying the frequency of ACC diagnosis over time and box and whiskers plot (10-90^th^ percentiles) representing the age at ACC onset. *compared to clusters 2, 3, and FS/NS/DEL. P-values on plots were calculated using Kruskal-Wallis tests. Mann-Whitney U tests were used for pairwise comparisons (*p<0.05, **p<0.01, ***p<0.001).

Upon analysis of cancer diagnoses in the NCI *TP53* Database, we noticed a drastic increase in frequency for cluster 5 carriers around age 30 due to a surge of breast tumor cases (**Fig. 4a**). Breast tumor onset varied considerably between the groups (p<0.0001, Kruskal-Wallis test), and again, carriers of clusters 3, 5, and FS/NS/DEL variants each had significantly earlier onset when compared to carriers of variants in clusters 1, 2, and 4 (**Fig. 4c** and **Supplementary Table 7**). Furthermore, in our analysis of ACC diagnosis, which is typically within the first few years of life, we found a significant difference between the patient groups (p=0.0095; Kruskal-Wallis test). In this case, cluster 4 variant carriers manifested ACC during young adulthood with a median age at diagnosis of 23 years, whereas all other patient groups experienced early childhood-onset ACC (**Fig. 4c** and **Supplementary Table 7**).

Finally, we investigated whether the *TP53* variant clusters could stratify the cancer survival of carriers. The Kaplan-Meier plot in **Figure 5a** shows that the lifetime cancer survival between cluster-based patient groups was significantly different (p<0.0001, log-rank test). Recapitulating the pattern of cancer onset, carriers of variants in clusters 1, 2, and 4 had considerably prolonged survival (median ages 74, 68, and 68, respectively) when compared to clusters 3, 5, and FS/NS/DEL carriers (median ages 52, 47, and 54, respectively) (**Supplementary Table 8**). The validation cohort demonstrated a similar trend in lifetime cancer survival (p=0.039, log-rank test) (**Fig. 5b**). Of note, the differences in observed survival time for cluster 5 when comparing the NCI *TP53* Database and validation dataset is likely due to monomeric p53 carriers. The validation cohort had only 1 monomeric variant carrier of 5 total cluster 5 carriers with survival data, whereas cluster 5 in the NCI *TP53* Database was comprised entirely of monomeric variant carriers (n=45). In addition, the observed shift in lifetime cancer survival between the NCI and validation datasets can be attributed to differences in data collection. The NCI *TP53* Database is a historical repository containing multigenerational data which provides lifelong survival projections. The validation dataset contains current patient records, some of which are pediatric, with less multigenerational data. As such, the validation cohort has more unaffected carriers with less lifetime data as compared to the NCI *TP53* Database (**Supplementary Table 9**), thus contributing to the discrepancy since the risk of death increases over time.

**Figure 5.**
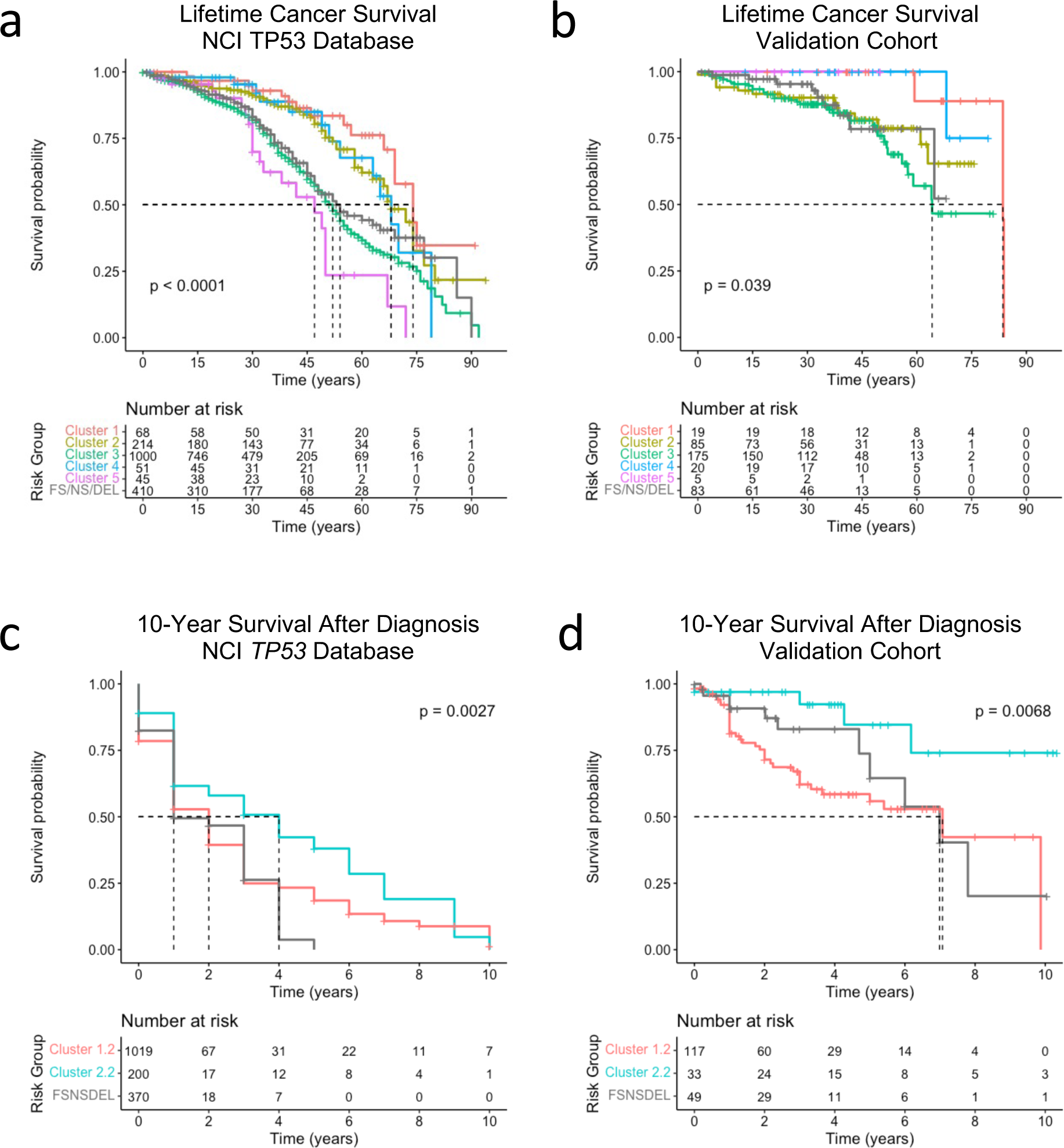
Survival analysis of *TP53* variant carriers. **(a)** Kaplan-Meier plots displaying the lifetime cancer survival of *TP53* variant carriers from the NCI *TP53* Database and **(b)** a validation cohort, stratified based on the variant clusters. **(c)** Kaplan-Meier plots showing 10-year survival time (age at cancer diagnosis to age at death) of *TP53* variant carriers from the NCI *TP53* Database and **(d)** a validation cohort, based on the 2-cluster model. p-values were calculated using the log-rank test.

In order to obtain adequate sample sizes with complete clinical data to analyze 10-year survival time after cancer diagnosis, we clustered patients into 2 groups (cluster 1.2 and 2.2) rather than 5 and kept FS/NS/DEL as an additional patient group (**Supplementary Fig. 4**). In this model, cluster 1.2 variants displayed greater structural and functional consequences as compared to cluster 2.2 variants (**Supplementary Figure 4d-g**). Remarkably, the cancer survival between these two patient groups along with FS/NS/DEL carriers was significantly different using data from the NCI *TP53* Database (p=0.0027, log-rank test) (**Fig. 5c**). Intriguingly, we observed an even greater stratification between survival outcomes when using the validation cohort (p=0.0068, log-rank test). In both cases, cluster 2.2 variant carriers had significantly extended cancer survival compared to cluster 1.2 (**Supplementary Table 10**). The differences in 10-year survival outcomes when using the NCI versus the validation cohort can be explained by the differences in cancer types between the datasets. Specifically, the validation cohort contained more breast tumors and fewer bone tumors when compared to the NCI *TP53* Database (**Supplementary Table 11**). Overall, this data suggests that the functionally distinct *TP53* variant clusters can stratify the cancer onset and survival of germline carriers, representing a promising risk stratification strategy for patients where there is currently none.

## DISCUSSION

The broad phenotypic spectrum related to *TP53* variants necessitates a risk stratification strategy. Diagnosis of LFS and the understanding of specific cancer risk is still a challenge because *TP53* variants are often rare with limited clinical evidence to base decisions on. The population prevalence of pathogenic variants is also unclear for the same reasons. Strikingly, *TP53* variants leading to protein changes occur in 7.8% of the general population (based on gnomAD v4.0, n=807,162 individuals; not including WT variant P72R), however most are likely benign. Current estimates suggest that pathogenic variants are found in 1 in 3555-5476 individuals [29]. This number was suggested to be as high as 1 in 400-865 using a less stringent definition of pathogenicity, although it is now considered an overestimate. Clarity around variant interpretations will help to avoid misclassification and improve patient management.

Here, we integrated functional screens using dimensionality reduction to gain a better understanding of the impact of different p53 variant species on their diverse cellular activities, including TA, DDR, LOF, and DN activity. Previous models to predict variant pathogenicity treated it as a binary variable and limited the focus to pathogenic versus benign [30, 31]. These studies did not examine the clinical effects of variants, whereas our approach has refined the categorization of pathogenicity and related these findings to clinical features to explore the spectrum of genetic variants. Using unsupervised clustering we identified 5 distinct *TP53* variant clusters with unique functional properties and clinical relevance. **Table 1** summarizes the main structural/functional features and clinical outcomes in each variant cluster, and **Supplementary Table 12** provides a reference for variant cluster assignments. Comparison of expert annotated variants with the clusters revealed a B/LB (cluster 1) and a P/LP (cluster 3) group, as well as three other groups with mixed pathogenicity interpretations or largely VUS (**Fig. 2a**). Since the majority of variants are not classified by VCEP and remain VUS, we used the clustering approach to classify all variants and found unique associations to cancer. Cluster 3 contained the most variants linked to cancer, and cluster 1 variants were the most prevalent in a non-cancer dataset (**Fig. 2b-e**). Moreover, cluster 1 variants were enriched in women >70 years who never had cancer.

**Table 1.** Summary of structural/functional features and clinical phenotypes associated with *TP53* variant clusters. *Data from monomeric variant carriers

| Variant Group | Structural Features and Regions | Functional Features | Defining Cancer Phenotypes | Cancer onset (median age in years with 95% CI) | Lifetime cancer survival (median age in years with 95% CI) |
| --- | --- | --- | --- | --- | --- |
| Cluster 1 | Disordered regions | Hypermorphic | Late onset/Less LFS core cancers | 42 (37-46) | 74 (69-NA) |
| Cluster 2 | DBD non-contact sites | Hypomorphic | – | 39 (35-41) | 68 (63-80) |
| Cluster 3 | DBD contact sites | LOF/DN | Very early onset breast cancer | 29 (28-30) | 52 (49-54) |
| Cluster 4 | CTD | WT-like function | Young adult ACC onset | 37 (31-47) | 68 (63-NA) |
| Cluster 5 | OD (monomeric) /TAD1/NTD hinge region | LOF/Hypomorphic | Osteosarcoma/Very early onset breast cancer* | 30 (28-33) | 47 (33-NA)* |
| FS/NS/DEL | Protein truncating | LOF | Very early onset breast cancer | 28 (27-30) | 54 (47-NA) |

We then tested the clinical impact by grouping carriers according to the clusters using discovery and validation cancer-associated datasets. Clear distinctions between groups, including unexpected findings, and consistent patterns emerged throughout our study. Specifically, a significantly higher frequency of osteosarcoma was discovered among cluster 5 carriers of monomeric p53 variants (**Fig. 3b**). Cluster 1 variant carriers presented with fewer LFS core cancers and a greater proportion being colorectal tumors (**Fig. 3c,d**). Furthermore, cancers in cluster 1 carriers were largely adult-onset and the lifetime survival in this group of patients was considerably extended compared to clusters 3, 5, and FS/NS/DEL groups (**Fig 4a,b** and **Fig. 5a,b**). In contrast, cluster 3 variants, which were primarily DBD variants with DN activity, predisposed to earlier cancer onset with shorter lifetime survival (**Fig. 4** and **Fig. 5a,b**). Cluster 2 variants were also mostly DBD variants but without DN activity, comprising a group of variants with intermediate pathogenicity compared to clusters 3 and 1. Cluster 4 carriers also displayed intermediate pathogenicity, and were distinguished by the delayed onset of ACC in young adulthood (**Fig. 4d**).

Taken together, the enrichment of cluster 1 variants in populations with no cancer history and the milder phenotypes associated with these carriers (i.e. older onset, superior lifetime survival, and less LFS-like tumor profiles) suggests that cluster 1 variants are B/LB. Their presence in the NCI *TP53* germline dataset may be a result of sampling bias. In fact, the relatively higher proportion of colorectal cancer in this variant cluster compared to other clusters reflects the proportion of worldwide cancer diagnoses that are colorectal cancer (10%) according to the World Health Organization. Previous work has suggested that multigene panel testing in colorectal cancer has expanded the mutational analysis of *TP53* to a wider range of patients, including those who do not meet classic LFS criteria, which can result in sampling bias that inflates cancer risk estimates [32].

A limitation of this study was the smaller sample size of the validation cohort that restricted the cancer type-specific analyses. Within the validation dataset, patient selection in the German cohort was more stringent for the inclusion of P/LP variants according to the Fortuno criteria [22], thus restricting the representation of variants in clusters outside of these criteria. In addition, the limited availability of clinical information made the analysis of survival after diagnosis less statistically rigorous when using 5 groups. However, by implementing a 2-cluster model to investigate 10-year survival, we found a remarkable separation between outcomes (**Fig. 5c,d**). In this model, cluster 1.2 variants encompassed the most severe functional consequences (including DN and monomeric variants) and led to significantly reduced survival time after diagnosis of cancer (**Supplementary Figure 4h** and **Supplementary Table 10**).

The current results suggest that a cancer risk continuum related to *TP53* variants extends beyond the LFS spectrum and can be risk stratified using integrative metrics (**Fig. 6**). At one end of the risk continuum, we find that cluster 1 variants retain higher functionality and are tolerable B/LB variants that were likely ascertained in association with cancer due to sampling bias (**Fig. 2d,e**). In contrast, we also identified high-risk variant carriers of clusters 3, 5, and FS/NS/DEL variants that result in earlier onset and poorer overall survival. The observation of reduced penetrance in clusters 2 and 4 may be the result of conditional variant pathogenicity under the influence of genetic or environmental modifying factors. Overall, the variant clusters provide a comprehensive new classification for variant interpretations and reveal novel risk factors that can be used to tailor an individual’s tumor surveillance plan and set clinical expectations. Considerations for modifying surveillance protocols according to the *TP53* variant clusters include (1) greater attention to bone lesions in whole-body MRI images from monomeric variant carriers; (2) intensive screening (particularly very early-onset breast cancer) for clusters 3, 5, and FS/NS/DEL carriers; (3) less intensive screening for clusters 2 and 4 carriers; (4) no screening recommended for cluster 1 carriers. Future development of this approach by incorporating more innovative mutagenesis screens may lead to even more refined risk stratification and personalized medicine.

**Figure 6.**
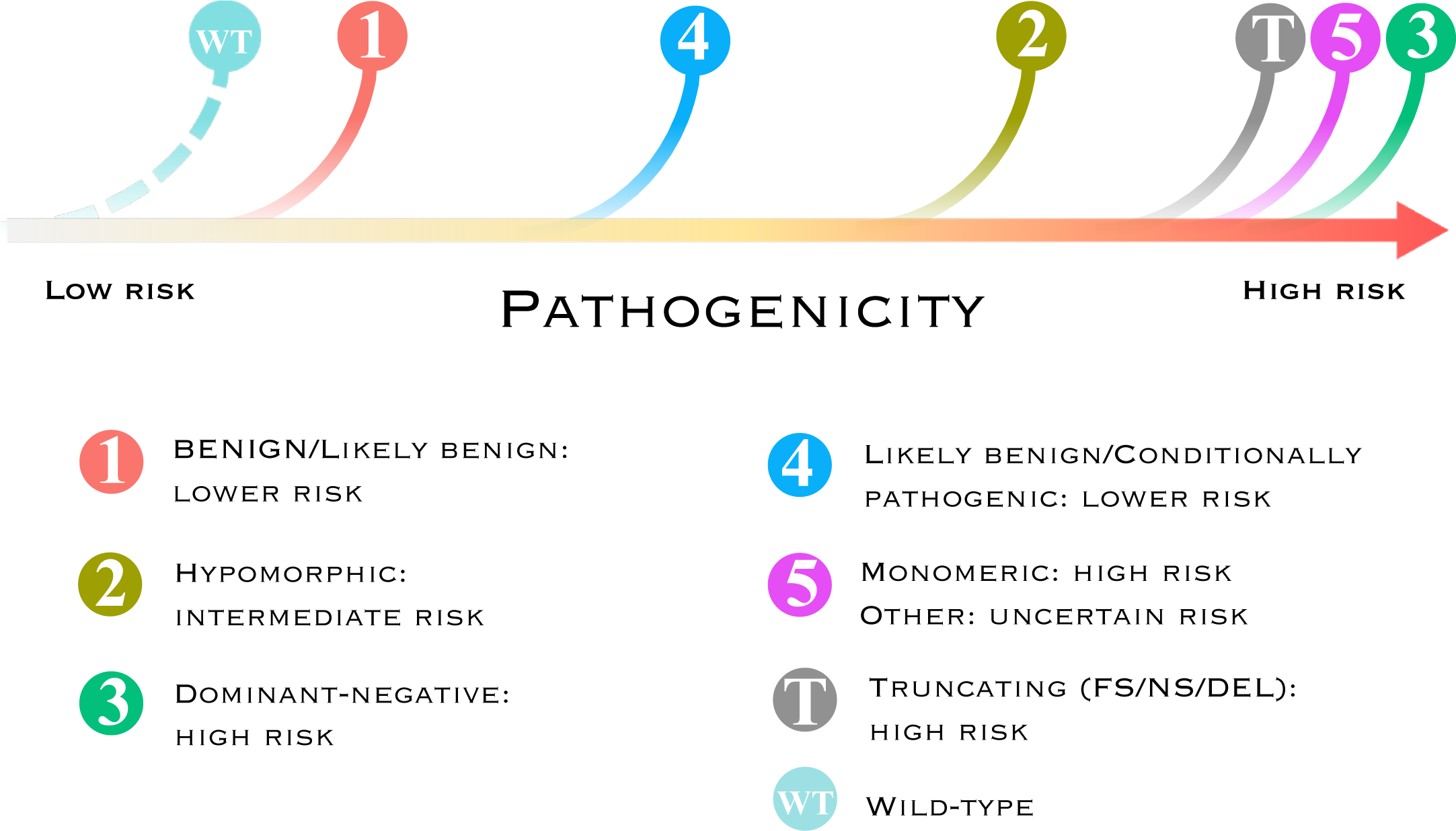
Interpretation of *TP53* variant clusters as a cancer susceptibility continuum based on cellular functional and clinical evidence.

## Data Availability

All data produced in the present work are contained in the manuscript

https://tp53.isb-cgc.org/

## ACKNOWLEDGEMENTS

This work is supported in part by a Terry Fox New Frontiers Program Project grant from the Terry Fox Research Institute (#1084). D.M. holds the CIBC Children’s Foundation Chair in Child Health Research. C.P.K. has been supported by the BMBF ADDRess (01GM2205A) and by the Deutsche Kinderkrebsstiftung (DKS2021.25).

