## Supplementary Figure 1 for "*TP53* Variant Clusters Stratify the Li-Fraumeni Spectrum and Reveal an Osteosarcoma-Prone Subgroup"

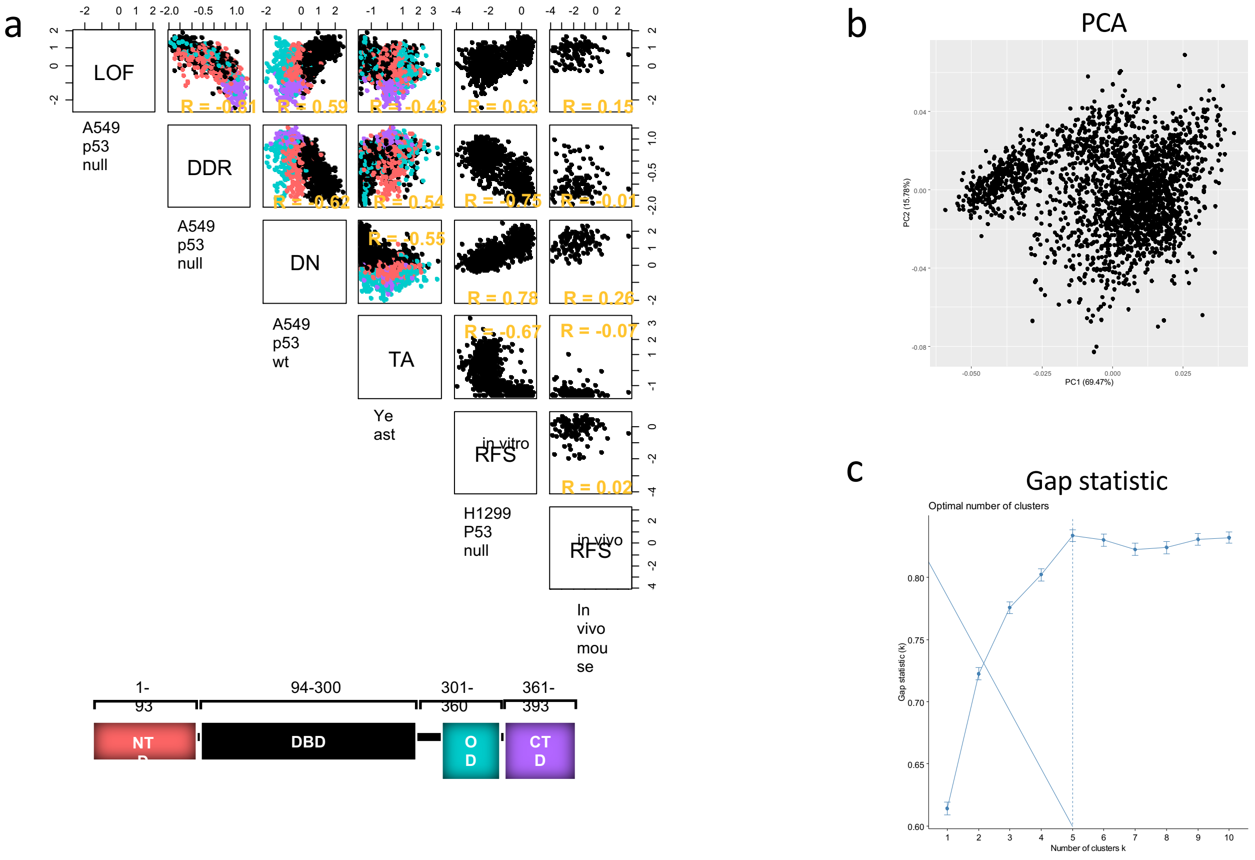


**Supplementary Figure 1. (a)** Correlation matrix plot comparing mutant p53 functional screens from mutagenesis libraries. **(b)** Principal component analysis (PCA) based on p53 functional features (LOF, DDR, DN effect, and TA). **(c)** Plot of the Gap statistical method plot to determine optimal number of clusters.
