## Supplementary Figure 2 for "*TP53* Variant Clusters Stratify the Li-Fraumeni Spectrum and Reveal an Osteosarcoma-Prone Subgroup"

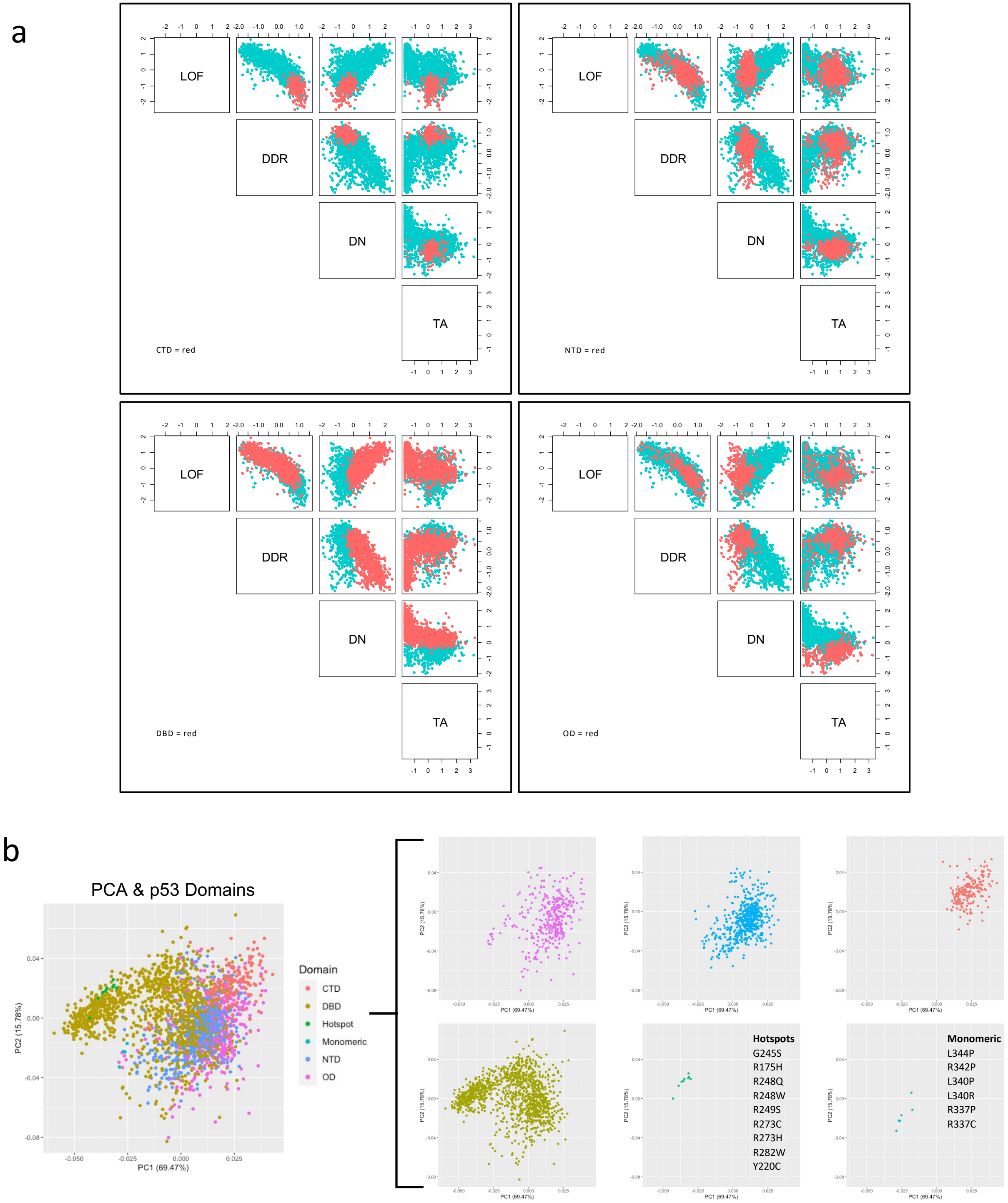


**Supplementary Figure 2.** Extended mutation resident domain analysis. **(a)** Matrix correlation plots comparing p53 mutagenesis screens, highlighting resident domain of interest in red. **(b)** PCA plot colour-coded based on the mutation resident domain
