## Supplementary Figure 3 for "*TP53* Variant Clusters Stratify the Li-Fraumeni Spectrum and Reveal an Osteosarcoma-Prone Subgroup"

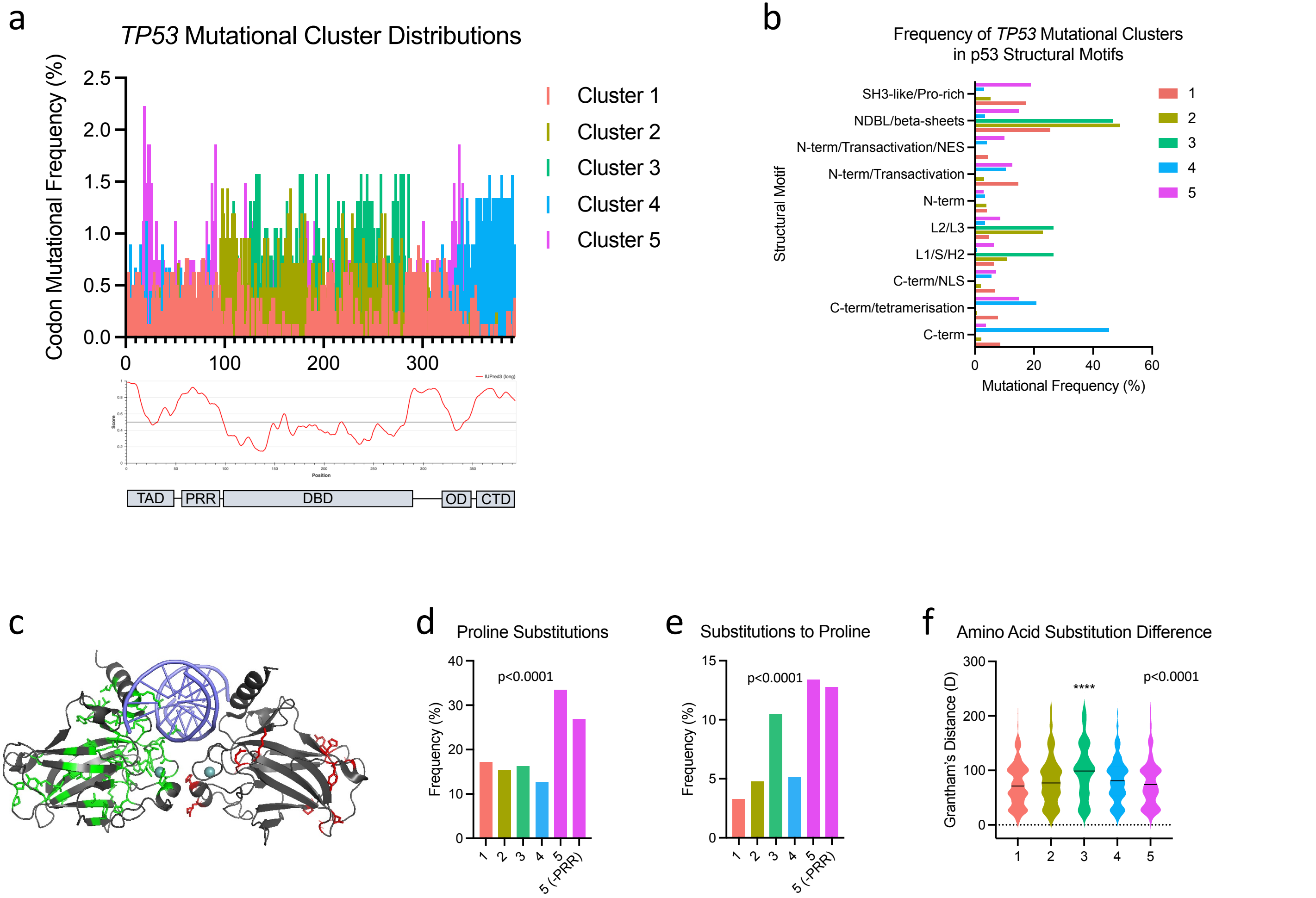


**Supplementary Figure 3.** Structure/function analysis of *TP53* variant clusters. **(a)** Codon distributions and frequencies of *TP53* variant clusters with intrinsically disordered regions shown below (determined using IUPred3 (Erdos 2021 Nucleic Acids Res)). **(b)** Bar graph showing the structural motifs where variant clusters reside. **(c)** Crystal structure of two p53 core DBDs bound to DNA (PDB #3EXJ) displaying the variant hotspots (cluster variant rate >1%) in clusters 2 and 3 (red and green, respectively). Side chains of the affected residues are shown, and DNA is represented in blue. **(d,e)** Frequency of proline residue substitutions. P-values were calculated using chi-squared tests. **(f)** Amino acid substitution differences as measured by Grantham's distance. P-value on plot was calculated using the Kruskal-Wallis test. Mann-Whitney U tests were used for pairwise comparisons (****p<0.0001).
