## Supplementary Figure 4 for "*TP53* Variant Clusters Stratify the Li-Fraumeni Spectrum and Reveal an Osteosarcoma-Prone Subgroup"

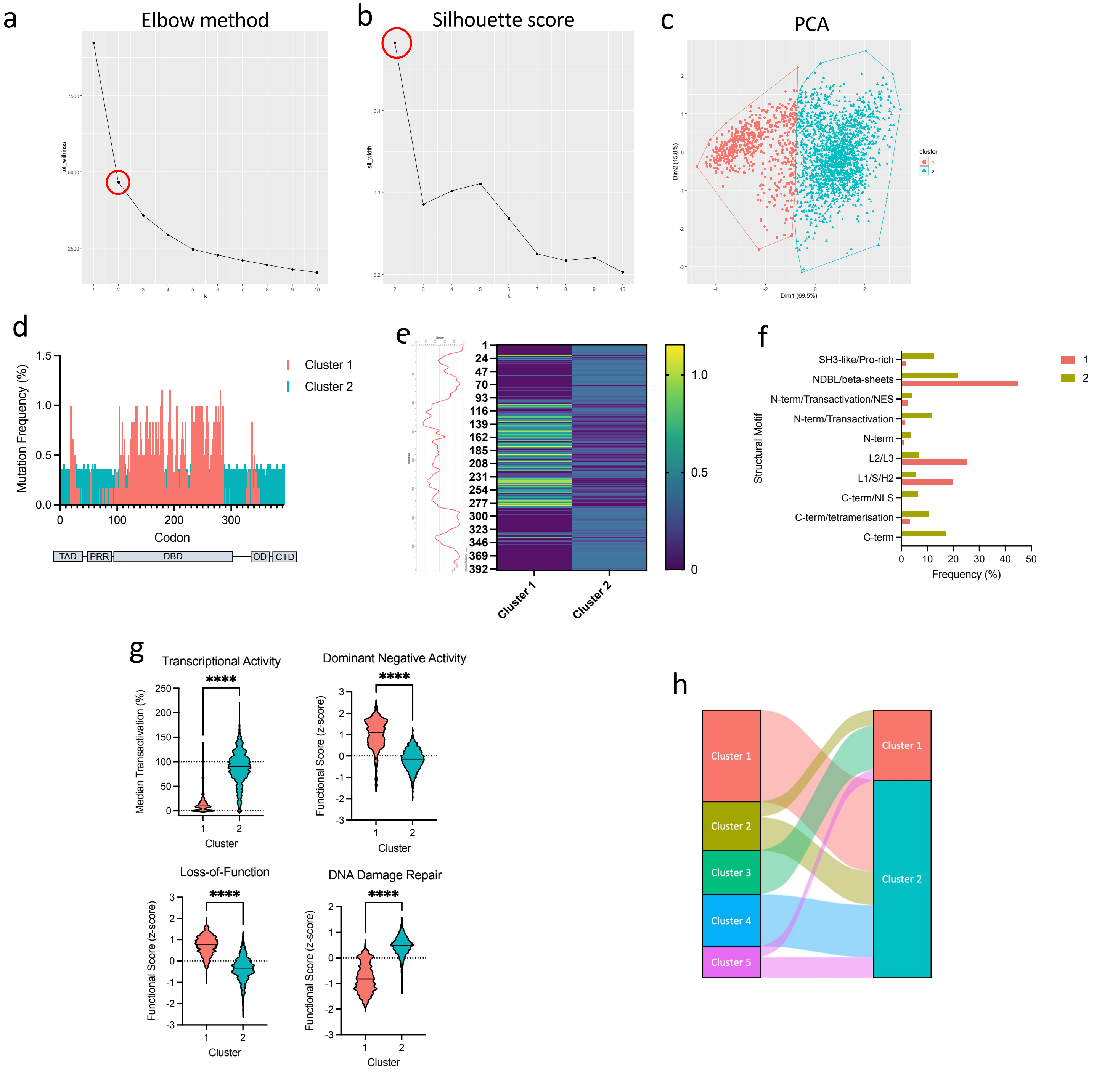


**Supplementary Figure 4.** *TP53* variant 2-cluster model analysis. **(a)** Elbow method plot and **(b)** Silhouette score plot to determine number of clusters. **(c)** Principal component analysis (PCA) unsupervised k-means clustering performed with p53 mutagenesis cellular functional assay measurements. **(d)** Codon distributions and frequencies of *TP53* variant clusters with intrinsically disordered regions shown below (determined using IUPred3 (Erdos 2021 Nucleic Acids Res)). **(e)** Heatmap displaying the codon frequencies and distributions of *TP53* variant clusters. **(f)** Bar graph showing the structural motifs where variant clusters reside. **(g)** Violin plots displaying the functional consequences of variants within each cluster (dotted line represents wild-type p53). ****p<0.0001, Mann-Whitney U test. **(h)** Alluvial plot comparing the variant clusters between the 2- and 5-cluster models.
